# Population-based reference equations and Z-scores for blood biomarkers of neurodegenerative diseases

**DOI:** 10.64898/2026.08.09.26360027

**Authors:** Sylvain Lehmann, Tahiana Andriambelosoa, Mehdi Morchikh, Marion Mortamais, Marie Duchiron, Xavier Ayrignac, Germain Busto, Christophe Hirtz, Audrey Gabelle, Karim Bennys, Sofiane Kab, Catherine Helmer, Marie Zins, Thibault Mura

## Abstract

**Background:** Blood biomarkers are increasingly used to support the diagnosis and monitoring of neurodegenerative diseases. However, their interpretation is complicated by physiological determinants, including age, sex, body-mass index, and renal function, and by differences in absolute concentrations between analytical methods. We aimed to develop population-based reference equations allowing individualized interpretation of selected blood biomarkers blood biomarkers used in neurology.

**Methods:** In this cross-sectional study, we analysed plasma samples from cognitively unimpaired participants selected from the French CONSTANCES and Three-City population-based cohorts. Generalized additive models for location, scale, and shape were used to model neurofilament light chain (NfL), glial fibrillary acidic protein (GFAP), phosphorylated tau 181 (p-tau181), amyloid-β40, amyloid-β42, and their ratios according to age, sex, body-mass index, and renal function. The resulting equations provided individualized expected concentrations, percentiles, and Z-scores. Previously established disease-specific concentrations were converted into Z-score. Cross-calibration equations were developed for NfL measurements across analytical methods and sample matrices.

**Findings:** The final reference populations comprised 5123 participants for amyloid biomarkers and p-tau181 and 5122 for NfL and GFAP; median age was 52.3 years and 49.7% were women. Between ages 40 and 80 years, expected NfL and GFAP concentrations increased by an average of 2.6% and 2.2% per year, respectively. Renal function, body-mass index, and sex had additional biomarker-specific effects. Application of the equations to clinical cohorts preserved distinct disease-associated profiles: NfL Z-scores were increased across disorders characterized by neuroaxonal injury, whereas p-tau181 and GFAP showed its greatest increase in Alzheimer disease. NfL cross-calibration equations showed excellent agreement between methods and matrices, with intraclass correlation coefficients greater than 0.90.

**Interpretation:** This population-based multibiomarker framework enables blood biomarker concentrations to be interpreted relative to individuals with similar physiological characteristics. Publicly available equations, reference curves, and standardized Z-scores could improve individualized interpretation and comparability across biomarkers, laboratories, and clinical populations.

## Background

Biological biomarkers represent a major asset in medical decision-making, as they represent quantifiable and reproducible elements contributing not only to the diagnosis of pathologies but also to risk assessment, prognosis, and therapeutic choice (1). Among biological fluids, blood is the most widely used and accessible medium for biological analysis, carrying a broad range of circulating proteins that reflect systemic physiological and pathological processes. In neurodegenerative and neuroinflammatory diseases, such as Alzheimer’s disease (AD), metabolites and proteins associated with pathological processes in the central nervous system (CNS) can cross the blood-brain barrier and reach the bloodstream, allowing their use as blood-based biomarkers.

The detection of CNS-derived biomarkers in blood has historically been challenging because their concentrations are substantially lower than those observed in cerebrospinal fluid (CSF), mainly due to dilution. The emergence of ultrasensitive approaches, such as SIMOA (2), has opened new avenues for the detection of CNS biomarkers in the blood. After an initial phase of validation, the demonstration that blood-based biomarkers could match the clinical performance of CSF biomarkers initiated a major transformation in the diagnosis of neurological diseases and was accompanied by new assays on clinical routine analyzers, whose sensitivity has been improved.

For example, the biological diagnosis of AD, which relies on the detection of pathologically relevant amyloid-beta (Aβ) peptides and phosphorylated tau (p-tau) proteins in the CSF, is rapidly shifting toward blood-based biomarkers (3–6). Detection of the astrocytic glial fibrillary acidic protein (GFAP), as a marker of astrocytic activation and damage, is moving to the blood (7–9). Importantly, plasma GFAP has shown stronger associations with AD-related amyloid pathology than CSF GFAP (10), whereas CSF GFAP has demonstrated limited diagnostic performance across dementia syndromes (11). Neurofilament light chain protein (NfL), which is released upon axonal damage of various origins, is another example of a blood-based biomarker (12, 13), with in vitro diagnostic (IVD)-certified methods now available in routine lab settings. Thus, we observed an exponential and rapid development in the use of these biomarkers for the biological diagnosis of neurological diseases.

The interpretation of blood-based biomarkers remains complex (12), as their values are affected by several physiological factors, such as age, sex, body mass index (BMI) and renal function (14). Thus, covariate-adjusted reference values are needed to guide interpretation, and to distinguish normal from abnormal changes. Several studies have developed these reference values, mostly for NfL and GFAP. For NfL, age has been consistently associated with higher concentrations (9, 15–20), while other studies also reported an inverse association with BMI (9, 20). Renal function may represent another important determinant, with lower estimated glomerular filtration rate (eGFR) associated with higher NfL concentrations reported (9). Similar age-related increase has been reported for GFAP, while lower eGFR, lower BMI, and female sex have also been associated with higher concentrations, even though the results are not consistent across studies (9, 15, 16). Importantly, most available reference data for NfL and GFAP were generated using Simoa-based assays, which may limit their applicability across analytical platforms.

In addition, reference values for other biomarkers are still limited, notably for AD-related biomarkers, including Aβ and p-tau. Indeed, their interpretation are now well-established in a pathological context. For example, in AD and related pathologies, Aβ42 and the Aβ42/Aβ40 ratio generally decrease, whereas p-tau tend to increase (18). However, among cognitively healthy participants without underlying pathologies, the change of these biomarkers is yet to be characterized. Cooper et al. reported that, among Canadian participants aged 3-79 years, the Aβ42/Aβ40 ratio showed a slight decrease with age, whereas p-tau181 followed a U-shaped association, with concentrations decreasing during childhood and increasing in later adulthood cooper (16). Accounting for other physiological determinants is needed for these biomarkers, as previous studies have shown that plasma levels of Aβ proteins and p-tau tend to increase with impaired renal function and decrease at higher levels of BMI (21, 22).

In this study, we developed reference values for a selection of relevant blood biomarkers of neurodegenerative diseases, including Aβ and p-tau proteins, NFL and GFAP using a large population-based cohort, measured with Roche Elecsys assay. We accounted for the effects of age, sex, BMI, and renal function, which are known to influence blood biomarker concentrations to varying degrees depending on the analyte (23). We also determined the optimal thresholds for normalized biomarkers using different IVD assays to detect AD, ALS, and MS. And finally, we provided cross-calibration equations to facilitate harmonization across different analytical platforms and biological fluids. The final goal is to provide the community with accessible equations and reference plots for direct clinical use, thereby improving the quality, interpretation, and performance of blood-based biomarkers for patient management in neurodegenerative and neurological disorders.

## METHODS

### Study design and populations

This cross-sectional study included participants from two French population-based cohorts, CONSTANCES and 3C. Ethical approval for the CONSTANCES study was obtained from the French Data Protection Authority (CNIL) and the INSERM Institutional Review Board, while the 3C study was approved by the Ethics Committee of the University Hospital of Kremlin-Bicêtre (No. 99-28). CONSTANCES is a nationwide cohort of over 200,000 adults aged 18-69 years at inclusion, randomly selected from the French National Pension Insurance Fund between 2012 and 2019 (24).The participants underwent standardized health examinations at 21 health screening centers and were followed up with periodic visits and annual questionnaires. Since 2018, biological samples have been collected and stored in the CONSTANCES biobank (25).

The 3C study enrolled 9,294 community-dwelling adults aged ≥ 65 years from Bordeaux, Dijon, and Montpellier between 1999 and 2001 (26). Participants underwent cognitive assessments, clinical examinations, and blood sampling at baseline and longitudinal follow-up.

Biomarker measurements were performed within the INTERVAL project in participants selected from two population-based cohorts: CONSTANCES, which includes more than 200,000 participants, and 3C. Given the logistical and financial constraints of measuring biomarkers in the entire cohorts, participants were randomly selected using stratified sampling based on age, sex, and BMI to ensure adequate representation across these major physiological determinants of biomarker concentrations. In CONSTANCES, 4,890 participants were included using 24 sampling strata, with additional inclusion of participants with impaired renal function (eGFR <60 mL/min per 1.73 m²) outside the stratified sampling to ensure adequate representation of this subgroup. In 3C, 483 participants were sampled across 36 strata. In CONSTANCES, a target of 200 participants was randomly selected per stratum, or all eligible participants when fewer than 200 were available, with additional inclusion of participants with impaired renal function. Consequently, selection probabilities varied across strata. Inverse probability weighting was applied to account for the sampling design and non-response.

For this analysis, we included all the participants from the INTERVAL project after excluding those with neurodegenerative diseases or substantial cognitive impairment (defined as a Mini-Mental State Examination (MMSE) score <24) at the time of blood sampling. This conventional threshold was chosen to exclude potentially pathological cognitive impairment while preserving the population-based nature of the reference sample. Covariates included age, sex (coded 1=female, 0=male), BMI, and markers of renal function (creatinine in µmol/L), estimated using the 2021 CKD-EPI creatinine equation (27).

### Datasets for clinical thresholds of normalized biomarkers

The ALZAN cohort, which was used to compute the AD thresholds, has been published (28). Briefly, it includes 423 participants who consulted memory clinics for cognitive complaints. The inclusion criteria required adults aged 50-85 years with cognitive complaints consistent with early stage AD. Participants were classified as amyloid-positive (Aβ+) or negative (Aβ-) based on their CSF Aβ42/Aβ40 (29). AD dementia was diagnosed according to the National Institute on Aging-Alzheimer’s Association (NIA-AA) criteria. Of the 423 ALZAN participants, 275 had complete data for age, sex, BMI, and creatinine and were therefore eligible for the fully adjusted Z-score analyses (Model 1). Patients were divided into two groups (ALS and non-ALS) according to their diagnosis based on the Clinical International Gold criteria (30). The retrospective MS cohort was constituted from the local MS clinic with 223 subjects, including primary progressive MS (PPMS) and relapsing-remitting MS (RRMS) patients, according to the 2017 McDonald criteria (31) and 28 neurological controls (NC). The demographic data are presented in Supplementary Table 3. Only patients with available age, sex, BMI, and creatinine levels were included to compute Z-scores. NfL and GFAP values were originally quantified in serum using Roche Elecsys and converted to plasma-equivalent values (see Supplementary Table 2). The Expanded Disability Status Scale (EDSS) was evaluated at withdrawal, and disease activity corresponded to either a recent relapse (<3 months) or new magnetic resonance imaging (MRI) lesions.

### Biochemical analysis

For biomarker measurement, blood aliquots were thawed at +4°C, gently homogenized, and centrifuged for 5 min at 2000 × g before analysis. One aliquot from each participant in each cohort was used for p-tau181, Aβ42, Aβ40, GFAP, and NfL measurements using Elecsys® assays on the e402 Cobas analyzer, as described elsewhere (32). Plasma measurements were performed across 56 analytical runs between April and November 2024, using the same reagent lot for each analyte and internal quality controls in each run. Between-run variability was evaluated as detailed in the Supplementary Methods. Intra- and inter-run coefficients of variation (CVs) were evaluated using plasma pools of clinical samples analyzed within a single run and across consecutive assay runs. For p-tau181, the intra- and inter-assay CVs were 2.5% and 3.7%, respectively, with an LLOQ of 0.30 pg/mL, respectively. For Aβ40, the corresponding values were 0.9%, 7.1%, and 10.0 pg/mL; for Aβ42, 3.2%, 4.5%, and 0.668 pg/mL; for GFAP, 1.63%, 2.93%, and 2.85 pg/mL; and for NfL, 1.92%, 9.70%, and 0.5 pg/mL. The AD, ALS and MS cohort samples were analysed using the same assays (3, 6, 7, 33). NfL and GFAP values were quantified in serum using Roche Elecsys and converted to plasma-equivalent values using cross-calibration, showing excellent agreement across platforms (ICC>0.9) (Supplementary Table 2).

### Impact of covariables

Relative covariate-related percentage changes were estimated using the fitted median (Z-score=0) biomarker values for women relative to men for each additional 1 year between 40 and 80 years, each unit increase in BMI between 20 and 40 kg/m², and 5 units increase in creatinine between 50 and 120 µmol/L. The 40–80-year interval was selected to provide a standardized summary of age-related changes across biomarkers. As age effects may be nonlinear, these average annual changes should not be interpreted as constant rates across the full age range covered by the reference curves.

### Statistical analyses

Population-based reference distributions were estimated using inverse probability weighted (IPW) generalized additive models for location, scale, and shape (GAMLSS) (34). These models allow the location, dispersion, skewness, and kurtosis of biomarker distributions to vary according to individual characteristics and are therefore well-suited for the derivation of age- and covariate-specific reference values.

Inverse probability weights were used to account for the sampling design and non-response. CONSTANCES participants received three weights (sampling cohort, sampling substudy, and nonresponse), whereas 3C participants received a rescaled weight based on national demographic distributions. The final weights were computed by multiplying the components.

### Four distributions

Box-Cox Cole Green (BCCG), Box-Cox T (BCT), Box-Cox power exponential (BCPE) and normal distributions were tested, with BCT providing the lowest Akaike Information Criterion (AIC) for all the biomarkers. BCT includes four distribution parameters: median µ, scale parameter σ (approximation of the coefficient of variation), skewness v, and kurtosis τ. Creatinine and eGFR were compared as alternative markers of renal function using the AIC. Creatinine-based models provided a better fit for all biomarkers except the Aβ42/Aβ40 ratio and were therefore retained in the final models to ensure consistency across biomarkers. Two sets of candidate covariates were considered for each biomarker: Model 1 included age, sex, BMI, and creatinine, whereas Model 2 considered only age and sex. Within each model, covariates were retained according to their contribution to model fit, as determined by the predefined selection procedure. For µ, covariates were added incrementally to the model and retained if they significantly reduced the AIC. Additionally, cohort effects were tested. For the selected continuous variables, linear and fractional polynomial terms (degrees 1 or 2) were tested. The same selection procedure for variables as for µ was applied for σ and v. To avoid overfitting, τ was not regressed on the covariates. The estimated distribution parameters (μ, σ, ν, and τ) were used to calculate the adjusted percentiles and Z-scores through a four-step procedure, allowing the interpretation of an individual concentration relative to the expected distribution in the reference population. Finally, age-specific reference curves were generated from the fitted models and plotted for selected Z-scores (0, 1, 1.5, and 2), stratified by creatinine concentrations (50, 80, and 100 µmol/L), BMI levels (20, 25, and 30 kg/m²), and sex. These values were selected for illustrative purposes: BMI values of 25 and 30 kg/m² correspond to the conventional thresholds for overweight and obesity, respectively, whereas creatinine concentrations of 50, 80, and 100 µmol/L represent different physiological profiles rather than established clinical cut-offs.

To evaluate the impact of physiological adjustment, we compared unadjusted Z-scores with Z-scores adjusted for age and sex (Model 2) and those additionally adjusted for BMI and creatinine (Model 1).. Linear regression models were then used to investigate the associations between diagnostic group and Z-score values for each approach. Disease-specific thresholds previously established in independent studies performed on the same analytical platforms were converted into Z-score thresholds using the reference equations developed in the present study.

Deming regression was performed to derive cross-calibration equations that allowed conversion between analytical platforms (35). This cross-calibration is important to convert biomarker measures performed on other fluids (serum vs. plasma) or with other analytical platforms before applying the equation. The Deming regression parameter λ was estimated using the ratio of x to y standard deviations (Sxx/Syy), where x and y correspond to those in the cross-calibration equations (36). The bootstrap method was used to estimate the 95% CI of the intercept and slope of the Deming regression equations. Agreement was assessed using intraclass coefficient (ICC) estimates and their 95% confidence intervals (CI) calculated based on single-rating, absolute agreement, and a 2-way random-effects model (ICC (2,1)) (37). Statistical analyses were performed using R 4.4.1 statistical software (R packages: gamlss, pROC).

Using these cross-calibration equations, the Z-score values were transformed to the Lumipulse IVD assay scale. Age-specific reference curves were then generated from the fitted models and plotted for selected Z-scores (0, 1, 1.5, and 2), stratified by creatinine concentrations (50, 80, and 100 µmol/L) for NfL levels. In addition, equations were used to calculate the Z-scores in the clinical cohorts. Linear regression models were fitted to assess the association between diagnosis and Z-scores. Finally, threshold values for the relevant biomarkers were estimated for each diagnosis by maximizing the Youden index.

## RESULTS

### Reference population

Of the 5,373 individuals initially considered, 134 were excluded at the time of blood sampling due to neurodegenerative diseases (n=64) or potential cognitive impairment (MMSE score <24; n=70). After further exclusions for missing data, values below the LLOQ, and outliers, the final reference populations comprised 5,123 and 5,122 cognitively unimpaired participants for amyloid biomarkers and p-tau, and for NfL and GFAP, respectively (Figure 1; Table 1). After IPW, the median age was 52.3 years (Q1–Q3, 42.3–64.0). The weighted age distributions of the two analytical populations are presented in Supplementary Figure 2. Both distributions showed the highest weighted frequencies in middle-aged participants, with lower frequencies at older ages.

**Figure 1.**
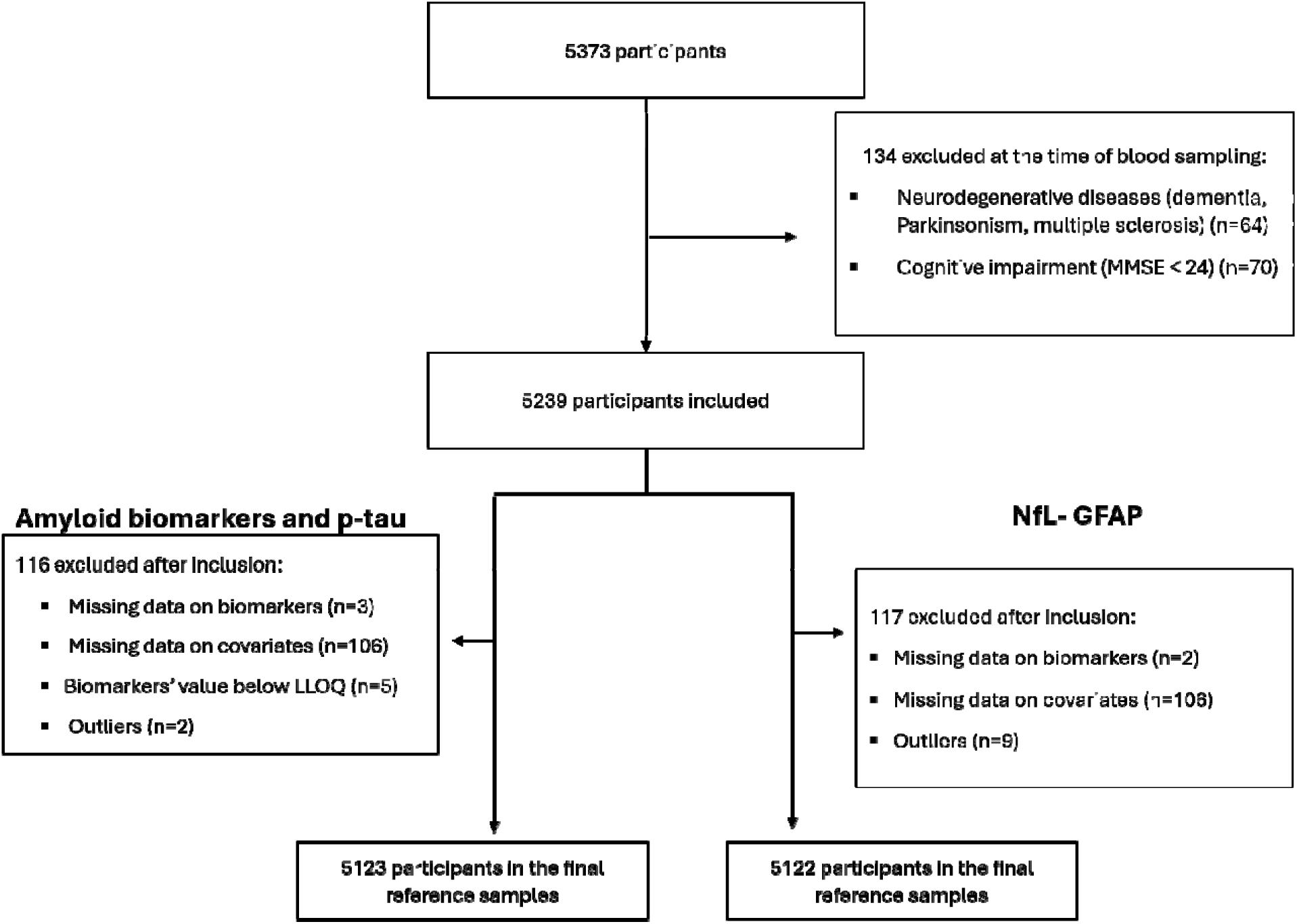
Flowchart of participant selection. Flowchart of participant selection. Participants were recruited from the French CONSTANCES cohort (n = 4,890) and the Three-City (3C) cohort (n = 483), for a total of 5,373 individuals. At the time of blood sampling, 134 participants were excluded because of neurodegenerative diseases (dementia, parkinsonism, or multiple sclerosis; n = 64) and cognitive impairment (MMSE < 24; n = 70). After inclusion, additional exclusions were applied because of missing biomarker measurements, missing covariate data, biomarker concentrations below the lower limit of quantification (LLOQ), and statistical outliers. The resulting reference populations differed according to the biomarker panel: 5,123 participants were included in the amyloid biomarker and p-tau analyses, whereas 5,122 participants were included in the NfL and GFAP analyses. **Abbreviations:** 3C, Three-City Study; LLOQ, lower limit of quantification; MMSE, Mini-Mental State Examination.

**Table 1:** Characteristics of the analytical reference populations used to derive the biomarker reference equations.

| **Characteristics** | **Amyloid and p-tau biomarkers**<br>**N=5,123** | **NfL-GFAP**<br>**N=5,122** |
|---|---|---|
| **Demographic characteristics** |  |  |
| **Age (years)** | 61.1 (46.3 ;70.6) | 61.1 (46.3 ;70.6) |
| **Sex, female** | 2541 (49.6) | 2545 (49.7) |
| **Clinical characteristics** |  |  |
| **BMI class (kg/m²)** |  |  |
| <25 | 1785 (34.8) | 1788 (34.9) |
| [25 ;30[ | 1813 (35.4) | 1808 (35.3) |
| ≥30 | 1525 (29.8) | 1526 (29.8) |
| **Impaired renal function** <sup>†</sup> | 261 (5.1) | 259 (5.1) |
| **Plasma biomarkers** |  |  |
| **Aβ40 (pg/mL)** | 241.0 (220.0;264.0) | 241.0 (220.0; 263.0) |
| **Aβ42 (pg/mL)** | 34.4 (30.8;38.0) | 34.4 (30.8; 38.0) |
| **p-tau181 (pg/mL)** | 0.7 (0.6;0.8) | 0.7 (0.6; 0.8) |
| **p-tau181/Aβ42 ratio** | 2.0 (1.7;2.5) | 2.0 (1.7; 2.5) |
| **Aβ42/Aβ40 ratio** | 14.4 (13.4;15.3) | 14.4 (13.4; 15.3) |
| **NfL (pg/ml)** | 1.9 (1.4; 2.6) | 1.9 (1.4; 2.6) |
| **GFAP (pg/ml)** | 67.1 (50.5; 91.1) | 67.0 (50.5; 91.0) |

The reference population comprised participants from the CONSTANCES and Three-City (3C) cohorts, selected through stratified sampling according to age, sex, and body mass index (BMI). Continuous variables are presented as median (first quartile [Q1]; third quartile [Q3]), and categorical variables as number (%).
† Impaired renal function was defined as an estimated glomerular filtration rate (eGFR) <60 mL/min/1.73 m². eGFR was calculated using the 2021 Chronic Kidney Disease Epidemiology Collaboration (CKD-EPI) creatinine equation.
**Abbreviations:** Aβ, amyloid-β; BMI, body mass index; GFAP, glial fibrillary acidic protein; NfL, neurofilament light chain; p-tau181, phosphorylated tau at threonine 181.

### Reference values for the different blood biomarkers

The reference equations are presented in Supplementary Table 1. In multivariable Model 1, age, sex, and renal function were retained as determinants for all biomarkers, whereas BMI was retained for all biomarkers except the p-tau181/Aβ42 ratio. p-Tau181 is used as an example in Table 2 to illustrate the four-step procedure for calculating p-tau181 z-scores. The influence of physiological covariates on the absolute biomarker concentrations corresponding to a given Z-score is further illustrated for NfL in Supplementary Figure 1.

**Table 2.**
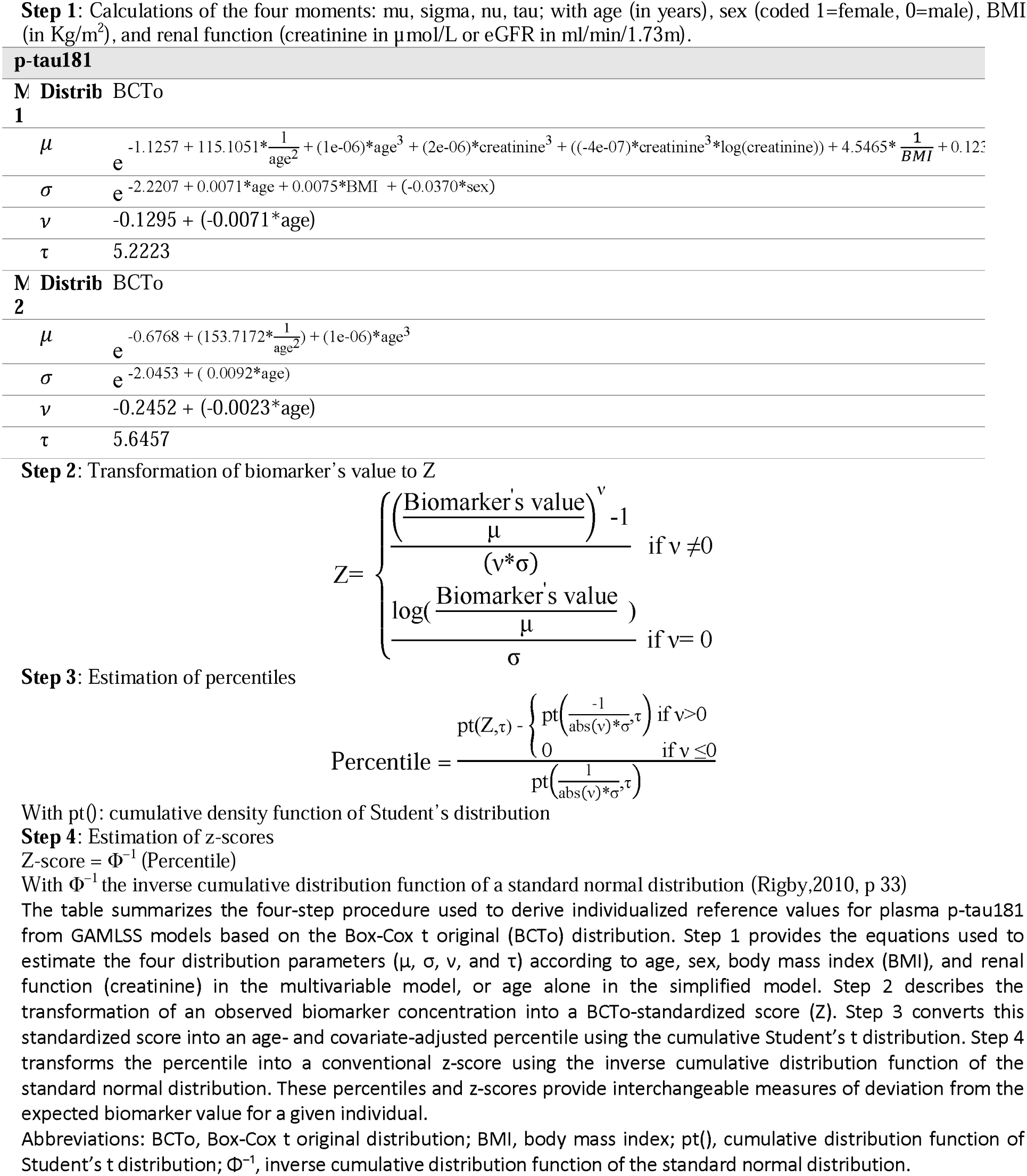
Equations and calculation procedure for deriving individualized percentiles and z-scores for plasma p-tau181.

| p-tau181 |  |  |
|---|---|---|
| M<br>1 | **Distrib** | BCTo |
|  | *μ* | $e^{-1.1257 + 115.1051*\frac{1}{age^{2}} + (1e-06)*age^{3} + (2e-06)*creatinine^{3} + ((-4e-07)*creatinine^{3}*log(creatinine)) + 4.5465*\frac{1}{BMI} + 0.123}$[illegible] |
|  | *σ* | e<sup>-2.2207 + 0.0071*age + 0.0075*BMI + (-0.0370*sex)</sup> |
|  | *ν* | -0.1295 + (-0.0071*age) |
|  | τ | 5.2223 |
| M<br>2 | **Distrib** | BCTo |
|  | *μ* | $e^{-0.6768 + (153.7172*\frac{1}{age^{2}}) + (1e-06)*age^{3}}$ |
|  | *σ* | e<sup>-2.0453 + ( 0.0092*age)</sup> |
|  | *ν* | -0.2452 + (-0.0023*age) |
|  | τ | 5.6457 |

**Step 2**: Transformation of biomarker’s value to Z
$Z= \begin{cases} \frac{\left(\frac{Biomarker's\ value}{\mu}\right)^{\nu}-1}{(\nu*\sigma)} & if\ \nu \neq 0 \\ \frac{log(\frac{Biomarker's\ value}{\mu})}{\sigma} & if\ \nu= 0 \end{cases}$
**Step 3**: Estimation of percentiles
$Percentile = \frac{pt(Z,\tau) - \begin{cases} pt\left(\frac{-1}{abs(\nu)*\sigma},\tau\right) & if\ \nu>0 \\ 0 & if\ \nu \leq 0 \end{cases}}{pt\left(\frac{1}{abs(\nu)*\sigma},\tau\right)}$
With pt(): cumulative density function of Student’s distribution
**Step 4**: Estimation of z-scores
Z-score = Φ<sup>−1</sup> (Percentile)
With Φ<sup>−1</sup> the inverse cumulative distribution function of a standard normal distribution (Rigby,2010, p 33)
The table summarizes the four-step procedure used to derive individualized reference values for plasma p-tau181 from GAMLSS models based on the Box-Cox t original (BCTo) distribution. Step 1 provides the equations used to estimate the four distribution parameters (μ, σ, ν, and τ) according to age, sex, body mass index (BMI), and renal function (creatinine) in the multivariable model, or age alone in the simplified model. Step 2 describes the transformation of an observed biomarker concentration into a BCTo-standardized score (Z). Step 3 converts this standardized score into an age- and covariate-adjusted percentile using the cumulative Student’s t distribution. Step 4 transforms the percentile into a conventional z-score using the inverse cumulative distribution function of the standard normal distribution. These percentiles and z-scores provide interchangeable measures of deviation from the expected biomarker value for a given individual.
Abbreviations: BCTo, Box-Cox t original distribution; BMI, body mass index; pt(), cumulative distribution function of Student’s t distribution; Φ<sup>−1</sup>, inverse cumulative distribution function of the standard normal distribution.

### Impact of covariates on biomarker concentrations

Figures 2, 3 and 4 present age-dependent reference curves for the selected biomarkers, illustrating the influence of creatinine, BMI, and sex on their expected concentrations. P-tau181, the p-tau181/Aβ42 ratio, NfL, and GFAP increased with age, with a steeper increase observed around 50-60 years of age (respectively Figure 2C, 2D, 2E, 2F). The largest increases were observed for NfL and GFAP, with concentrations rising by 2.6% and 2.2% per year, respectively, between 40 and 80 years (Table 3). The Aβ42/Aβ40 ratio gradually declined with age (Figure 2B), with a 0.2% decrease per year. For Aβ42, only modest age-related changes were observed (+0.3% per year) (Figure 2A). The age-dependent effects of physiological covariates on NfL are further illustrated in Supplementary Figures 1.

**Figure 2.**
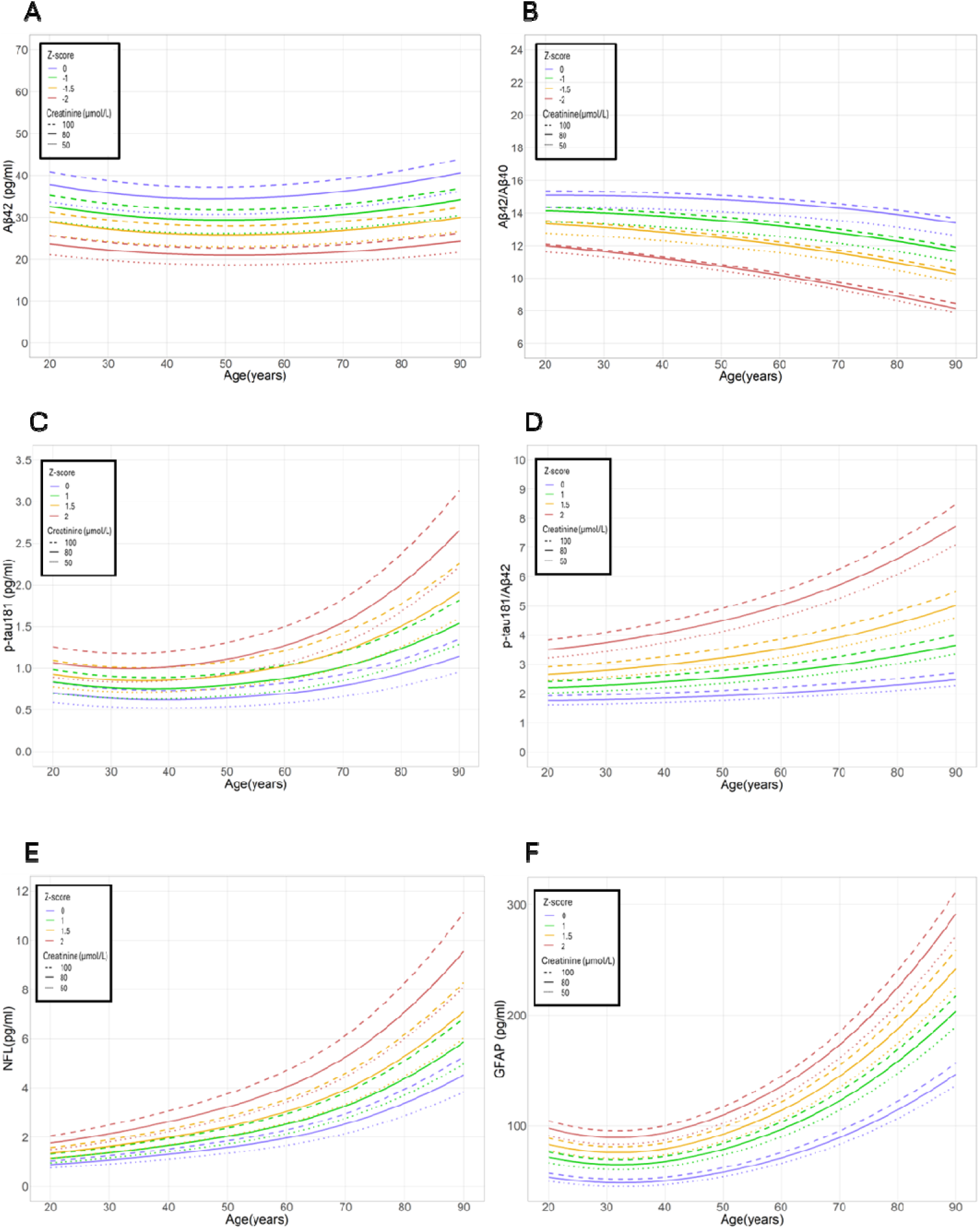
Reference curves illustrating the effect of renal function. Reference curves were derived from the final GAMLSS models and represent biomarker concentrations according to age for selected Z-scores (0, ±1, ±1.5, and ±2; colored lines). Panels A-F show Aβ42 (A), Aβ42/Aβ40 ratio (B), p-tau181 (C), p-tau181/Aβ42 ratio (D), NfL (E), and GFAP (F). Dashed, solid, and dotted lines correspond to creatinine concentrations of 100, 80, and 50 µmol/L, respectively. Negative Z-scores are shown for Aβ42 and Aβ42/Aβ40, and positive Z-scores for the other biomarkers. **Abbreviations:** GAMLSS, generalized additive models for location, scale, and shape; NfL, neurofilament light chain; GFAP, glial fibrillary acidic protein.

**Figure 3.**
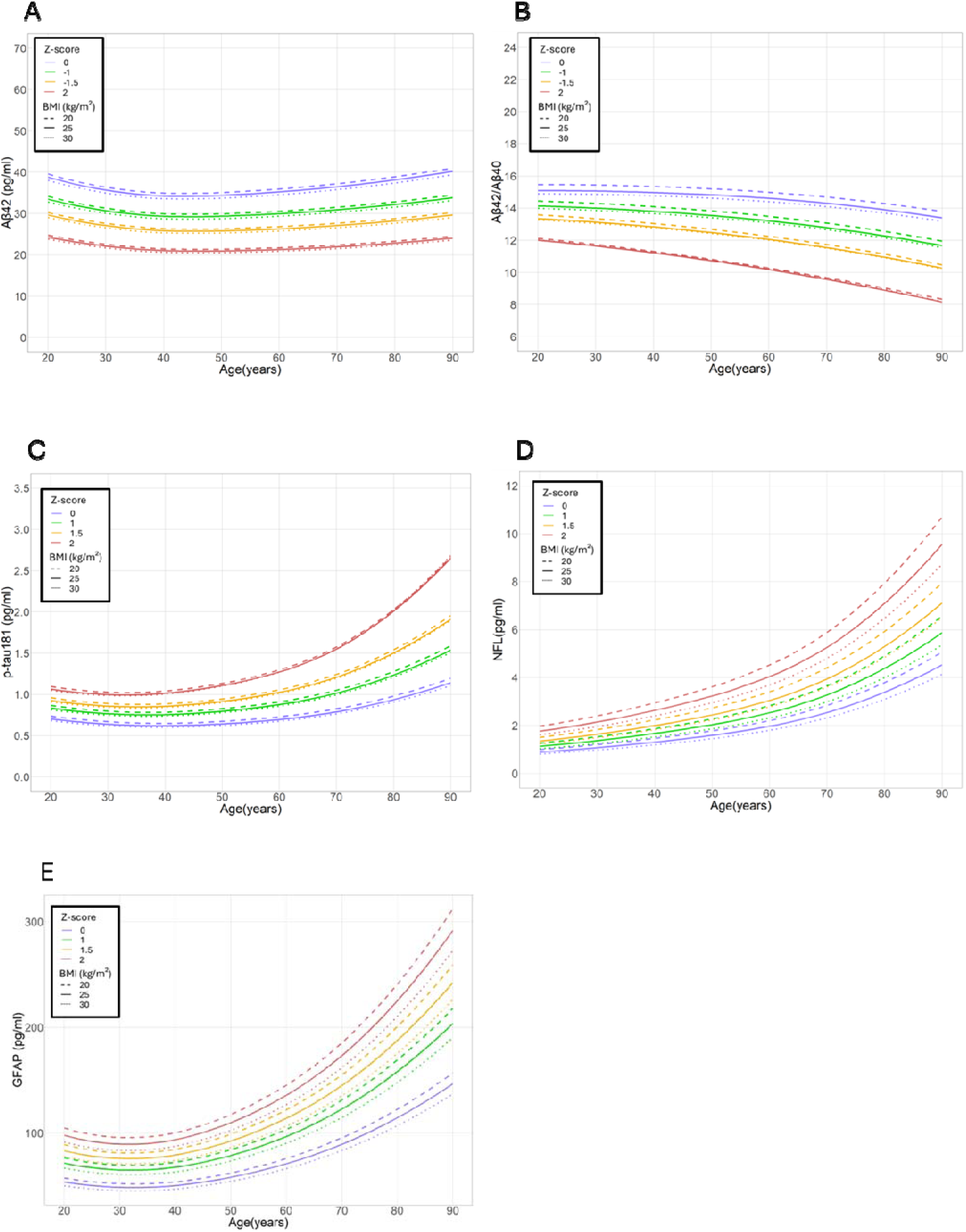
Reference curves illustrating the effect of body mass index. Reference curves were derived from the final GAMLSS models and represent biomarker concentrations according to age for selected Z-scores (0, ±1, ±1.5, and ±2; colored lines). Panels A-E show Aβ42 (A), Aβ42/Aβ40 ratio (B), p-tau181 (C), NfL (D), and GFAP (E). Dashed, solid, and dotted lines correspond to BMI values of 20, 25, and 30 kg/m², respectively. Negative Z-scores are shown for Aβ42 and Aβ42/Aβ40, and positive Z-scores for the other biomarkers. **Abbreviations:** BMI, body mass index; GAMLSS, generalized additive models for location, scale, and shape; NfL, neurofilament light chain; GFAP, glial fibrillary acidic protein.

**Figure 4.**
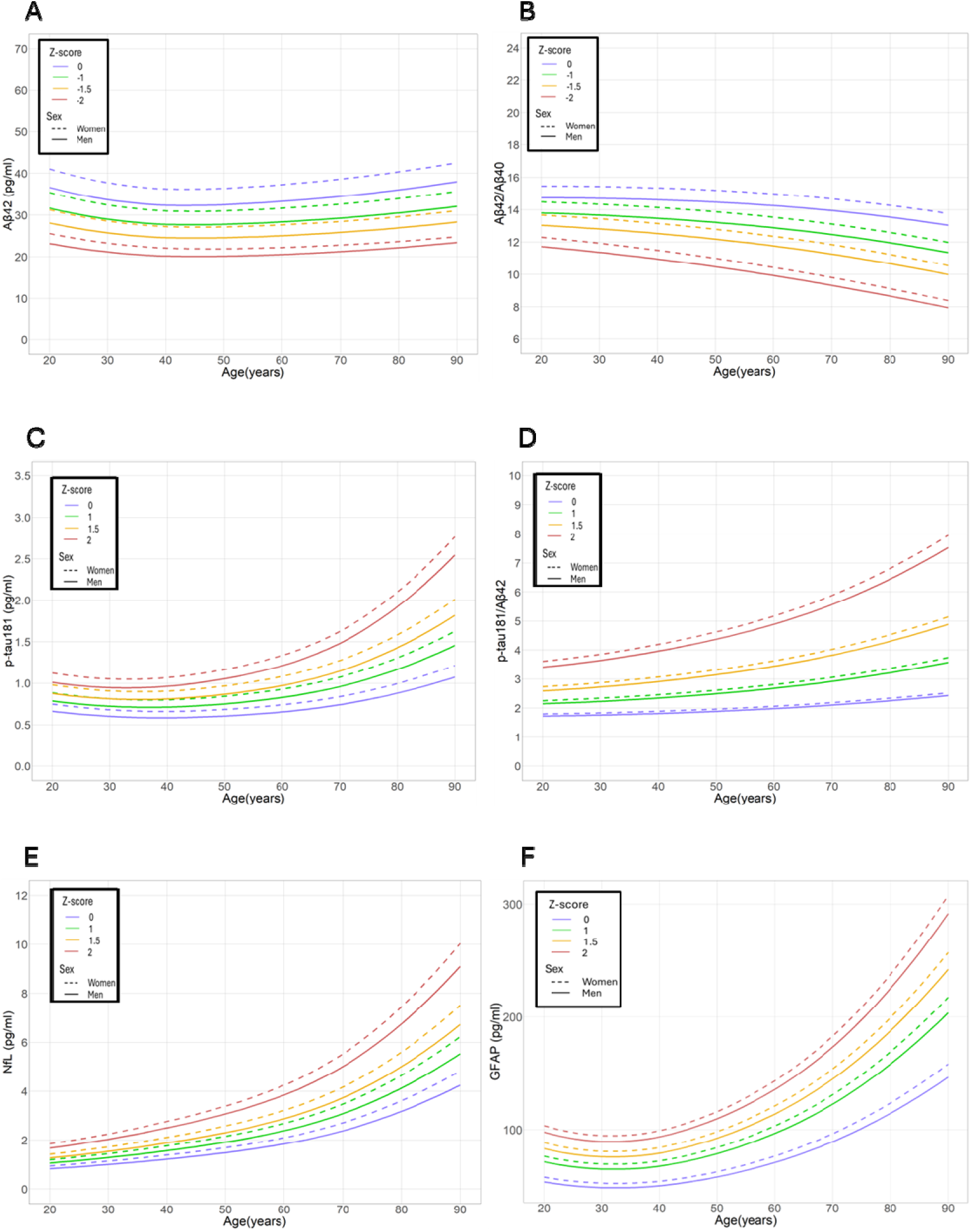
Reference curves illustrating the effect of sex. Reference curves were derived from the final GAMLSS models and represent biomarker concentrations according to age for selected Z-scores (0, ±1, ±1.5, and ±2; colored lines). Panels A-F show Aβ42 (A), Aβ42/Aβ40 ratio (B), p-tau181 (C), p-tau181/Aβ42 ratio (D), NfL (E), and GFAP (F). Dashed and solid lines correspond to women and men, respectively. Negative Z-scores are shown for Aβ42 and Aβ42/Aβ40, and positive Z-scores for the other biomarkers. **Abbreviations:** GAMLSS, generalized additive models for location, scale, and shape; NfL, neurofilament light chain; GFAP, glial fibrillary acidic protein.

**Table 3:** Relative effects of age, renal function, BMI, and sex on plasma biomarker concentrations.

| **Biomarker** | **Age (1 year)** | **Creatinine (5 µmol/L)** | **BMI (1 kg/m<sup>2</sup>)** | **Women vs Men** |
|---|---|---|---|---|
| **NfL** | +2.6% | +3.6% | -1.6% | +4.3% |
| **GFAP** | +2.2% | +1.6% | -1.3% | +16.6% |
| **Aβ42** | +0.3% | +2.0% | -0.4% | +12.0% |
| **Aβ42/Aβ40** | -0.2% | +0.6% | -0.3% | +5.1% |
| **p-tau181** | +1.0% | +3.9% | -0.6% | +13.2% |
| **p-tau181/Aβ42** | +0.6% | +2.0% | – | +4.0% |

**Legend:** Values show the average relative percentage change in biomarker concentration associated with the following increases, estimated across the specified ranges:
- 1 year of age (ages 40–80 years)
- 5 µmol/L increase in serum creatinine (50–120 µmol/L)
- 1 kg/m<sup>2</sup> increase in BMI (20–40 kg/m<sup>2</sup>)
- Female sex relative to male sex

Regarding creatinine effects, higher creatinine levels were associated with higher concentrations across all biomarkers (Figure 2), with the largest effects observed for p-tau181 and NfL (+3.9% and +3.6%, respectively, per 5 µmol/L increase in creatinine). For BMI, an increased BMI was generally associated with lower biomarker concentrations (Figure 3), particularly for NfL (-1.6%) and GFAP (-1.3%) (Table 3), whereas its effect was more limited for amyloid biomarkers and p-tau181. Female sex was associated with higher expected concentrations across all biomarkers (Figure 4), with the strongest effects observed for GFAP (+16.6%), p-tau181 (+13.2%), and Aβ42 (+12.0%) (Table 3).

### Clinical application of the reference equations

After excluding participants with missing data on age, sex, creatinine, or BMI, the characteristics of the patients included in the independent clinical cohorts are summarized in Supplementary Table 3. The Alzan cohort included 275 patients (median age, 74.2 years [IQR 68.8-78.1]; 60.4% women), of whom 216 (78.5%) had AD and 23 (8.4%) had FTD. The ALS cohort comprised 150 patients (median age, 68.3 [63.7-74.6] years; 58.7% women), including 100 (66.7%) patients with ALS. The MS cohort included 210 patients after exclusion of individuals younger than 23 years (median age 48.0 years [35.0-55.0]; 76.7% women), including 150 (71.4%) with relapsing-remitting MS (RRMS) and 34 (16.2%) with primary-progressive MS (PPMS).

To evaluate the clinical relevance of the proposed reference equations, biomarker concentrations were converted into age- and covariate-adjusted Z-scores. Figure 5 compares disease–control differences for GFAP, NfL, and p-tau181 using unadjusted Z-scores, Model 2, and Model 1. Overall, disease-associated differences were largely preserved across the three approaches, although physiological adjustment modified their magnitude without consistently increasing disease-control separation. For GFAP, the highest values were observed in AD participants compared to controls (β = 1.28, 95% CI 0.89-1.67 with the model 1), while PPMS and FTD exhibited more moderate elevations of GFAP Z-scores. In contrast, p-tau181 Z-scores were selectively increased in AD (β = 1.95, 95% CI 1.59-2.32 with the model 1). For NfL, Z-scores values were consistently elevated across all diagnostic groups except RRMS, with the largest increases observed in FTD (β = 1.61, 95% CI: 1.06-2.17 with the model 1), consistent with widespread neuroaxonal injury. The magnitude of the estimated differences was generally slightly reduced within the adjusted models, although the overall pattern of biomarker alterations across diagnostic groups remained largely consistent across the three models.

**Figure 5.**
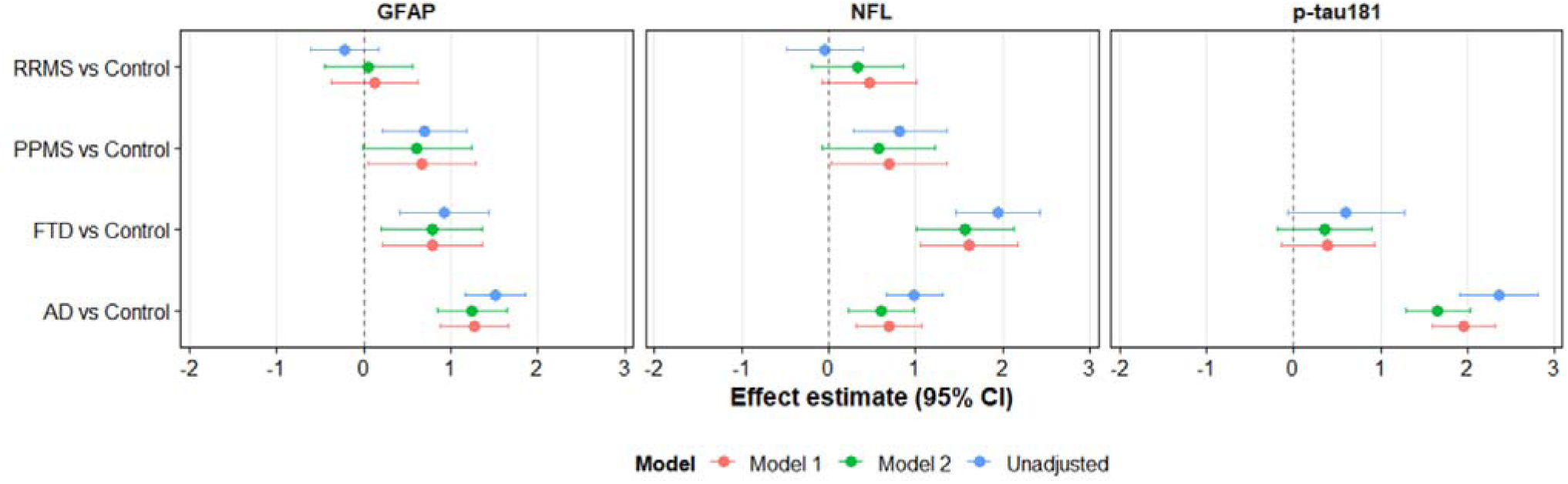
Comparison of unadjusted and covariate-adjusted plasma biomarker Z-scores across clinical cohorts. Estimated differences in biomarker Z-scores between patients and controls for Alzheimer’s disease (AD), frontotemporal dementia (FTD), primary progressive multiple sclerosis (PPMS), and relapsing-remitting multiple sclerosis (RRMS). Panels show GFAP, NfL, and p-tau181. Three approaches were compared: unadjusted Z-scores (blue), Z-scores adjusted for age and sex (Model 2, green), and Z-scores additionally adjusted for BMI and creatinine (Model 1, red). Points represent regression coefficients (β), expressed as differences in Z-score units between each disease group and its corresponding controls, and horizontal bars indicate 95% confidence intervals. The vertical dashed line at 0 indicates no difference between patients and controls. **Abbreviations:** AD, Alzheimer’s disease; BMI, body mass index; FTD, frontotemporal dementia; GFAP, glial fibrillary acidic protein; NfL, neurofilament light chain; PPMS, primary progressive multiple sclerosis; RRMS, relapsing-remitting multiple sclerosis.

The disease-specific optimal Z-score thresholds are summarized in Table 4. Consistent with the observed biomarker profiles, the Aβ42/Aβ40 ratio showed negative cut-offs in AD (-1.40) and FTD (-1.23), reflecting lower-than-expected values relative to the reference population. The highest NfL thresholds were observed in ALS (1.86) and FTD (1.53) patients.

### Cross-calibration equations

The cross-calibration equations are presented in Supplementary Table 2, showing excellent agreement between the evaluated analytical platforms and sample matrices (ICC >0.90). These equations allow biomarker concentrations measured using alternative assays or in serum to be converted to the Roche plasma-equivalent scale used to establish our reference distributions. This enables the calculation of comparable Z-scores across the evaluated methods and matrices, facilitating the interpretation of results from different clinical cohorts, studies, and laboratories.

### Web platform

The normative values derived from this study are available through an interactive R Shiny application, allowing for the comparison of an individual’s biomarker concentrations with those of the reference population. The application is freely accessible at: https://insinter.shinyapps.io/appli/.

## DISCUSSION

In this study, we established population-based reference values for five individual blood biomarkers of neurodegenerative diseases, including NfL, GFAP, Aβ40, Aβ42, p-tau181, and two biomarker ratios, Aβ42/Aβ40 and p-tau181/Aβ42. Using a large reference population comprising more than 5,000 cognitively unimpaired individuals’ representative of the French population, we developed percentiles and Z-scores adjusted for major physiological determinants. To facilitate implementation in clinical and research settings, we provide both mathematical equations and graphical reference curves, allowing the direct conversion of biomarker concentrations into standardized scores. Their clinical relevance has been illustrated in independent cohorts, including AD, FTD, ALS, and MS. Importantly, although physiological adjustment modifies disease–control differences, this does not necessarily translate into improved diagnostic accuracy, which was not directly assessed in this comparison.

Population-based reference studies have increasingly extended beyond NfL. Cooper et al. established age-specific reference intervals for Aβ42/Aβ40, p-tau181, NfL, and GFAP (16), while Rousset et al. developed multivariable reference intervals for NfL and GFAP (9). More recently, the large HUNT study characterized age-related plasma p-tau217 distributions and the prevalence of biomarker-defined AD pathology in 11,486 participants aged 58 years and older (38). Direct numerical comparisons of reference limits across studies remain challenging because of differences in analytical platforms, sample matrices, population characteristics, and covariate adjustment strategies. Our study complements these investigations by providing covariate-adjusted reference equations and individualized Z-scores for five blood biomarkers and two ratios measured using Roche Elecsys assays.

The covariate effects observed in our study were consistent with those reported in previous studies of biomarker reference values, which showed that NfL (9, 15–20), GFAP (9, 15, 16), p-tau181 (16) levels increased with age and with impaired levels of renal function marker and lower BMI. In addition, the annual percentage changes estimated in our study were within the range previously reported in the literature, with increases of approximately 3-4% per year for NfL (16, 19), 2% for GFAP (16), and approximately 3% for p-tau181 among individuals aged 60 years and older (16). We also observed consistently higher levels of these biomarkers among women compared with men. Our findings are consistent with those reported by Rousset et al. for GFAP, although other studies have not observed a significant association between sex and GFAP levels. This discrepancy may be attributable to differences in study populations, age distributions, analytical platforms, or other unmeasured biological or methodological factors. Physiological factors influence circulating biomarker concentrations to markedly different extents. NfL and GFAP, which are non-specific biomarkers to neurodegenerative disease, are the most influenced by age, while creatinine has strong influence on NfL and p-tau181. The inverse associations with BMI may partly reflect a blood-volume dilution effect, as previously proposed for NfL (14). Age-related changes may partly reflect alterations in biomarker clearance (39). The association with age remained significant, even after adjusting for renal function. However, renal function was assessed using creatinine, which is an imperfect surrogate of kidney function, particularly in older individuals, where creatinine levels are strongly influenced by muscle mass (40). Recent studies have highlighted the important influence of kidney function on blood-based neurological biomarkers (23), and our study confirmed and quantified this effect. However, the influence of renal function on biomarker concentrations does not necessarily translate into improved diagnostic accuracy after adjustment. Arslan et al. (41) reported that adding eGFR to models including plasma biomarkers, age, and sex did not improve amyloid-positivity discrimination, although their cohort predominantly included individuals with preserved or mildly impaired renal function. Sex-related differences were also observed across most biomarkers, with women generally exhibiting higher expected concentrations than men after adjusting for other covariates, particularly for GFAP, NfL, and p-tau181. Previous population-based reference studies have reported inconsistent sex effects ranging from no detectable differences across biomarkers (16), to higher GFAP concentrations in women (9), suggesting that these associations may depend on the population studied and the biomarker considered. The biological basis for this difference remains incompletely understood. The persistence of a sex effect after adjustment for BMI and renal function suggests that sex captures additional biological determinants that are not fully explained by these variables. Population-specific differences in cumulative behavioural and health-related exposures may also contribute to the heterogeneity observed across studies.Consequently, sex appears to be an important component of individualized biomarker interpretations.

The influence of age, renal function, BMI, and sex was not uniform across all biomarkers. Aβ peptides, which have both central and peripheral origins, were less influenced by BMI. Notably, Aβ42/Aβ40 and p-tau181/Aβ42 were less strongly influenced by these physiological determinants. This likely reflects the fact that both analytes composing the ratio are influenced in similar ways by these physiological factors, such that calculating the ratio tends to offset their effects (42–44).

The establishment of normative equations made it possible to express biomarker concentrations as standardized Z-scores rather than relying solely on absolute concentrations. Standardized Z-scores reduce physiological variability attributable to age, sex, BMI, and renal function, thereby improving the comparability across individuals. In addition, because all biomarkers are expressed in standard deviation units, Z-scores enable direct comparison of abnormalities across biomarkers, despite their different analytical scales and concentration ranges. Similar to pediatric growth charts or bone mineral density Z-scores, this framework expresses biomarker concentrations relative to the expected distribution for an individual with similar physiological characteristics. Z-scores also facilitate longitudinal monitoring by enabling changes to be interpreted relative to the expected biological variation. This normalization may improve diagnostic and prognostic assessments by distinguishing disease-related biomarker abnormalities from physiological variations.

Individualized reference equations may be useful in several clinical settings. First, they may facilitate the interpretation of biomarker results in routine laboratory practice by providing standardized measures of abnormalities. Second, they may improve harmonization across clinical centers by reducing the variability attributable to differences in patient characteristics. Third, they may support patient selection, risk stratification, and treatment monitoring in the clinical trials. This approach may be especially valuable in older populations, where physiological increases in biomarker concentrations complicate the interpretation of the absolute values. The potential clinical benefit of these reference equations is likely biomarker-dependent. For NfL and GFAP, which are relatively nonspecific and strongly influenced by physiological factors, adjusted Z-scores may help identify elevations beyond those expected for an individual’s physiological profile. In contrast, for AD-related biomarkers such as p-tau181, disease-associated changes may substantially exceed physiological variability, and established assay-specific diagnostic cut-offs validated against amyloid PET or CSF biomarkers should remain the primary basis for diagnostic classification. Adjusted Z-scores may provide complementary information, particularly for borderline values or patients with relevant comorbidities, but their additional diagnostic value remains to be established. Such individualized interpretation aligns with the broader movement toward precision medicine and may facilitate the integration of blood biomarkers into routine neurological care in the future.

Disease-associated biomarker profiles remained largely consistent after adjustment for physiological covariates, without clear evidence of improved diagnostic discrimination. Nevertheless, adjusted Z-scores provide a standardized measure of biomarker deviations relative to the expected distribution for individuals with comparable physiological characteristics. Benkert et al. (20) demonstrated that age- and BMI-adjusted serum NfL Z-scores improved the identification of disease activity and prognostication in MS compared with absolute concentration cutoffs. In our study, we found that accounting for creatinine and BMI may slightly attenuate NfL association with MS clinical phenotypes, underlying that part of the differences may be due to underlying comorbidities. However, overall patterns are consistent. Rather than masking disease-related biological differences, Z-scores enable biomarker abnormalities to be interpreted relative to the expected value for an individual with a similar age, sex, BMI, and renal function. However, physiological normalization does not confer disease specificity. Even biomarkers considered relatively specific for AD, such as p-tau181 and p-tau217, may be elevated in other neurological disorders, including Creutzfeldt– Jakob disease (45) and amyotrophic lateral sclerosis (46). As blood biomarkers are generally measured in patients with a clinical suspicion of neurological disease, Z-scores should be interpreted in the context of the clinical presentation and complementary diagnostic investigations, rather than considered as standalone evidence of AD.

In addition to individualized interpretation, this study also provides assay conversion equations between major analytical platforms, including Elecsys®, Lumipulse®, Ella®, and Simoa® assays. Although blood biomarkers are increasingly used in both research and clinical settings, substantial inter-method variability remains a major obstacle to their implementation. Absolute concentrations may differ considerably between assays, despite excellent correlations. Combined with assay conversion equations, expressing biomarker concentrations as percentiles or Z-scores relative to a common reference population provides a unified interpretative framework across the analytical platforms. Such harmonization could facilitate the comparison of results between studies, laboratories, and healthcare systems. Standardized reference values may contribute to future international efforts aimed at biomarker standardization and regulatory implementation.

This study had several important strengths. First, it is based on a large reference population exceeding 5,000 participants, making it one of the largest normative studies of blood neurodegeneration biomarkers to date. Another strength of this study is the simultaneous evaluation of several clinically relevant biomarkers using a unified statistical framework. Finally, the use of GAMLSS modeling allowed us to estimate not only the mean biomarker values but also the age-dependent changes in variability, skewness, and kurtosis, providing a more flexible representation of biomarker distributions than conventional linear approaches.

### Limitation

This study has several limitations. First, the study population was predominantly composed of French individuals of European ancestry, and external validation in more diverse populations is necessary. Second, the analyses were based on cross-sectional data and therefore, cannot fully characterize within-subject biomarker trajectories over time. Third, only a single biomarker measurement was available for each participant, preventing the assessment of biological and analytical variability at the individual level. Fourthly, cognitive impairment was defined using an MMSE cut-off score of 24, a threshold intended to exclude substantial cognitive impairment while preserving the population-based nature of the reference sample. Consequently, some participants with subtle cognitive impairment may have remained in the reference population.

In addition, relatively few participants were older than 85 years, which limited the precision of the reference estimates at the extreme upper end of the age distribution. Future studies should evaluate the performance of these equations in independent cohorts and assess their impact on diagnostic accuracy and clinical decision-making. The same methodological framework could also be extended to emerging biomarkers such as p-tau217, brain derived (BD) p-tau217, endogenously cleaved tau species from the microtubule-binding region (eMTBR), α-synuclein, and other markers of neurodegeneration and neuroinflammation. The integration of these equations into laboratory information systems, web-based calculators, and automated reporting tools may further facilitate their adoption in routine clinical practice.

### Conclusions

In conclusion, we developed population-based reference values and standardized Z-scores for the principal blood biomarkers of neurodegenerative diseases. By accounting for age, sex, BMI, and renal function, this framework enables the individualized interpretation of biomarker concentrations and addresses important sources of physiological variability. The proposed framework may facilitate harmonized reporting across laboratories and analytical platforms and improve clinical interpretation by expressing biomarker abnormalities on a common standardized scale, enabling direct comparisons across patients, neurological diseases, and biomarkers.

## Data Availability

Requests will be considered by each study investigator based on the information provided by the requester regarding the study and analysis plan. Individual-level data from the CONSTANCES cohort cannot be made publicly available because they contain sensitive health information and are protected by French data protection regulations. Access may be granted to qualified researchers following the submission and approval of a research proposal, completion of the applicable legal and regulatory procedures, and establishment of an agreement with the CONSTANCES cohort. Information on the application procedure is available on the CONSTANCES scientific access portal.

---

**Supplementary Figure 1.**
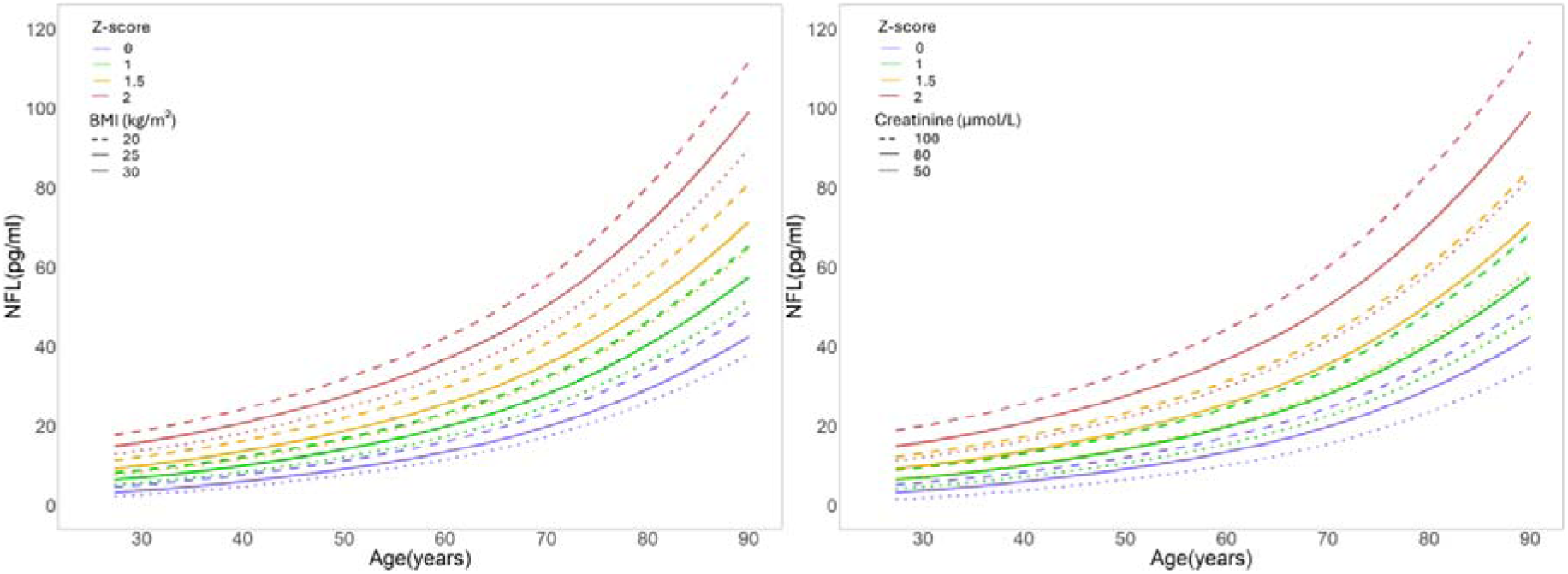
Influence of age, body mass index, and renal function on plasma NfL reference concentrations and Z-scores. Model-predicted plasma NfL concentrations (pg/mL) are plotted as a function of age for Z-scores of 0 (purple), 1 (green), 1.5 (orange), and 2 (red), based on the multivariable reference equations. The left panel illustrates the influence of body mass index (BMI: 20, 25, and 30 kg/m²), while the right panel illustrates the influence of creatinine concentrations (50, 80, and 100 µmol/L). Line styles distinguish BMI or creatinine values, as indicated in each panel. These curves illustrate how physiological covariates influence the absolute NfL concentrations corresponding to a given Z-score.

**Supplementary Figure 2.**
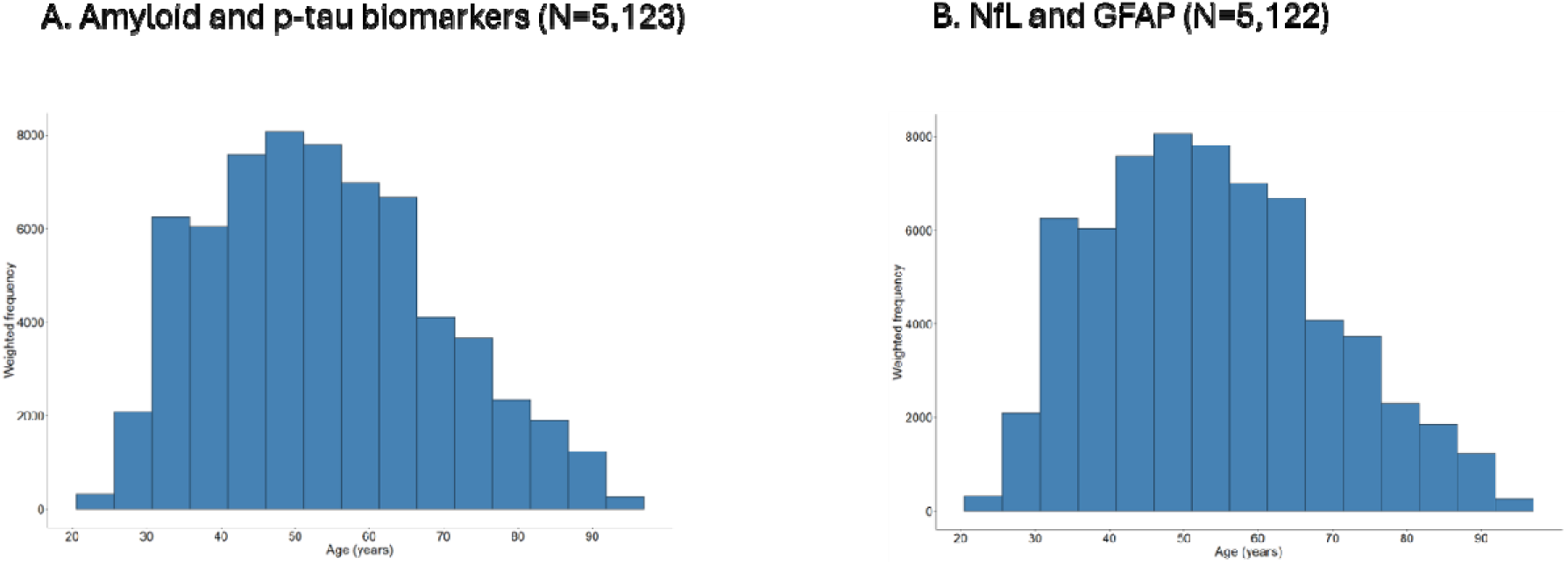
Age distribution of the reference population. Age distributions of participants included in the reference analyses for (A) amyloid and p-tau biomarkers (n=5,123) and (B) NfL and GFAP (n=5,122). Histograms represent inverse probability-weighted frequencies to account for the sampling design and non-response. Weighted frequencies do not correspond to the actual numbers of participants.

## Abbreviations

3C: Three-City Study
Aβ: amyloid beta
Aβ40: amyloid beta 40
Aβ42: amyloid beta 42
AD: Alzheimer’s disease
AIC: Akaike information criterion
ALS: amyotrophic lateral sclerosis
BCCG: Box-Cox Cole-Green
BCPE: Box-Cox power exponential
BCT: Box-Cox t
BMI: body mass index
CI: confidence interval
CNS: central nervous system
CSF: cerebrospinal fluid
CV: coefficient of variation
EDSS: Expanded Disability Status Scale
eGFR: estimated glomerular filtration rate
FTD: frontotemporal dementia
GAMLSS: generalized additive models for location, scale, and shape
GFAP: glial fibrillary acidic protein
ICC: intraclass correlation coefficient
IPW: inverse probability weighting
IVD: in vitro diagnostic
LLOQ: lower limit of quantification
LP: lumbar puncture
MMSE: Mini-Mental State Examination
MRI: magnetic resonance imaging
MS: multiple sclerosis
NfL: neurofilament light chain
NIA-AA: National Institute on Aging-Alzheimer’s Association
PPMS: primary progressive multiple sclerosis
p-tau181: phosphorylated tau 181
RRMS: relapsing-remitting multiple sclerosis.

## Declarations

## Ethics approval and consent to participate

The CONSTANCES cohort study was approved by the Institutional Review Board (IRB) of the National Institute of Health and Medical Research (Inserm) (Opinions No. 01-011 and No. 21-842) and authorized by the French data protection authority (CNIL) No. 910486; Biobank was approved by the CPP Sud Est I (Opinion No. 2018-32) and CNIL Authorization No. DR-2-2018-137. The 3C study was approved by the Ethics Committee of the University Hospital of Kremlin-Bicêtre (No. 99-28). Written informed consent was obtained from all participants.

## Consent for publication

non applicable

## Availability of data and materials

The reference equations, model coefficients, and calculation procedures required to estimate the expected biomarker concentrations, Z-scores, and percentiles are provided in the article and are publicly available. Automated Excel spreadsheets for calculating individual Z-scores and percentiles are available from the corresponding author.

## Competing interests

Consultant or Advisory Role: S Lehmann, Advisory Board for Roche Diagnostics, Biogen, Lilly, and Fujirabio. A Gabelle, Advisory Board member for Biogen, Lilly, and Esai.

## Funding

AXA Mécénat Santé (INTERVAL Project), Fondation Recherche Alzheimer (ALZAN project), Fondation pour la Recherche Médicale (FRM, team Proteinopathies), Caisse nationale d’assurance maladie, and other institutional partners of CONSTANCES and 3C studies.

The funders had no role in the study design, data collection, data analysis, data interpretation, or writing of the report. The corresponding author had full access to all data and had the final responsibility for the decision to submit the manuscript for publication.

## Authors’ contributions

**TM:** Drafting/revision of the manuscript for content, including medical writing for content; study concept or design; analysis or interpretation of data. **TA:** Drafting/revising the manuscript for content, including medical writing for content; analysis or interpretation of data. **MeM, MaM, MD, KB, and SK:** drafting/revision of the manuscript for content, including medical writing for content, and analysis or interpretation of data. **AG, MZ, and CH:** drafting/revision of the manuscript for content, including medical writing for content; analysis or interpretation of data; major role in the acquisition of data. **SL:** Drafting/revision of the manuscript for content, including medical writing for content; major role in the acquisition of data; study concept or design; analysis or interpretation of data; has full access to all the data in the study and take the final responsibility for the decision to submit for publication.

## Declaration of generative AI and AI-assisted technologies in the manuscript preparation process

During the preparation of this work, the authors used Gemini and ChatGPT, provided within their professional working environment, to assist with table formatting, verification of the consistency of table and figure citations, and English-language editing. The authors reviewed and edited all outputs as needed and take full responsibility for the content of the published article.

## Acknowledgement

The Constances Cohort Study was supported and funded by the Caisse Nationale d’ Assurance Maladie (CNAM). The Constances Cohort Study is an Infrastructure nationale en Biologie et Santé and benefits from a grant from ANR (ANR-11-INBS-0002) and the Ministry of Research. Constances is also partly funded by MSD, AstraZeneca, and Lundbeck. AXA Mécénat Santé (INTERVAL Project) for the population-based study, Fondation Research Alzheimer (ALZAN projet) for the clinical cohort. The 3C Study was conducted under a partnership agreement between the Institut National de la Santé et de la Recherche Médicale (INSERM), Victor-Segalen Bordeaux-2 University, and Sanofi-Aventis. The Fondation pour la Recherche Médicale supported the preparation and initiation of the study. The 3C study was also supported by the Caisse Nationale Maladie des Travailleurs Salaries; Direction Générale de la Santé; MGEN; the Institut de la Longevité; Agence Nationale de la Recherche ANR PNRA 2006 (06-01-01) and Longvie 2007 (LVIE-003-01); Agence Française de Sécurité Sanitaire des Produits de Santé; the Regional Governments of Aquitaine, Bourgogne, and Languedoc-Roussillon; the Fondation de France; the Ministry of Research-INSERM Programme Cohorts and collection of biological material; Fondation Plan Alzheimer” (FCS 2009-2012); the Caisse Nationale de Solidarité pour l’Autonomie (CNSA); Roche Pharma; and the Agence nationale de sécurité sanitaire de l’alimentation, de l’environnement et du travail (Anses, grant N◦ 2019/1/116).

## Notes

### Author Declarations

The CONSTANCES cohort study was approved by the Institutional Review Board (IRB) of the National Institute of Health and Medical Research (Inserm) (Opinions No. 01-011 and No. 21-842) and authorized by the French data protection authority (CNIL) No. 910486; Biobank was approved by the CPP Sud Est I (Opinion No. 2018-32) and CNIL Authorization No. DR-2-2018-137. The 3C study was approved by the Ethics Committee of the University Hospital of Kremlin-Bicetre (No. 99-28). Written informed consent was obtained from all participants.

### Summary of Updates

Following the request for revision we have improve the manuscript and in particular the figures that were not optimal for reading. We also added information on methodology and citations but we did not change data or conclusions.

